# Comfort with AI for HIV Prevention Among Cisgender Women in New York City

**DOI:** 10.64898/2026.06.02.26354471

**Authors:** Harry Reyes Nieva, Max Flanagan, Simian Huang, Deborah A. Theodore, Amelie Foumena Nkodo, Melissa Parkinson, Sarah Hill, Megan McAndrew, Jorge Benitez, Henry Peralta, Silvia Amesty, Jason E. Zucker, Magdalena E. Sobieszczyk, Delivette Castor

## Abstract

Long-acting pre-exposure prophylaxis (PrEP) expands HIV prevention options for women. However, PrEP impact depends on addressing persistent gaps in awareness, access, and use. Artificial intelligence (AI) tools, including conversational agents, are being explored to advance PrEP uptake, but comfort with AI may influence their impact. Thus, we examined women’s comfort with AI and its association with PrEP awareness. We analyzed self-reported data from women aged *≥*18 years in a cross-sectional survey conducted in New York City from August 2023 to August 2024. We performed descriptive analyses, applied latent class analysis to identify AI knowledge/comfort profiles, and estimated unadjusted and adjusted odds ratios to assess associations between profile membership and PrEP awareness. Among 306 respondents without a diagnosis of HIV who completed AI-related survey items, the median age was 36. Most women identified as Hispanic/Latina (60%) or Non-Hispanic Black (18%), had not completed college (53%), and spoke only English or were bilingual (81%). Latent class analysis identified four AI knowledge/comfort profiles that differed by PrEP awareness, race/ethnicity, borough, prior drug use, and technology utilization. Women with varied AI knowledge, broad AI discomfort, and comfort with clinicians maintaining privacy had lower odds of PrEP awareness (OR: 0.35, 95% CI: 0.16-0.75), but this association did not persist after statistical adjustment. PrEP awareness and AI knowledge were limited, yet many women expressed openness to AI-enabled tools when privacy was assured. AI-enabled HIV prevention tools should prioritize trust, transparency, confidentiality, and the lived contexts of the women they intend to serve.

## Introduction

Women assigned female at birth account for nearly 20% of new HIV infections in the United States (US). HIV pre-exposure prophylaxis (PrEP) is highly effective in preventing HIV acquisition, yet PrEP awareness, access, and uptake remain suboptimal among US women, with roughly 10% indicated for PrEP receiving a prescription.^1,2^ Barriers to oral PrEP use are multilevel, including structural factors such as stigma, medical mistrust, and limited access to high-quality sexual healthcare; clinical factors such as inconsistent counseling and prescribing practices; and individual-level factors including low perceived HIV risk, confidentiality concerns, and preference for newer methods such as long-acting PrEP.^3–9^ As HIV PrEP methods expand, person-centered strategies for education and engagement are increasingly important to translate biomedical advances into clinical and public health impact.

Digital health interventions, including artificial intelligence (AI)-enabled conversational agents and decision support systems, are increasingly being explored as implementation strategies to extend HIV prevention education, risk assessment, screening, counseling and linkage to care. AI-enabled tools have supported HIV testing navigation, PrEP counseling and education, risk assessment, and care engagement.^10–18^ AI and other computer-mediated tools may facilitate disclosure of sensitive information about sexual practices, substance use, mental health, intimate partner violence, and HIV risk by offering more privacy and reducing perceived judgment or embarrassment. Therefore, AI-enabled tools could bolster PrEP impact through tailored, private, and scalable support for HIV prevention.

As societal adoption has increased, willingness to use AI-enabled tools has varied substantially by context and public trust remains mixed.^19^ People generally view AI more favorably when it supports administrative, informational, or assistive tasks than when it is used for judgment-intensive functions such as diagnosis, treatment, decision-making, or psychological counseling. Prior research suggests that perceived usefulness, performance expectancy, effort expectancy, attitudes, and trust influence adoption of AI-enabled technologies.^20^ Trust generally refers to the belief that a system will perform reliably, appropriately, and safely under conditions of uncertainty and vulnerability.^20^ Comfort with AI is related but distinct, reflecting one’s sense of ease with using, interacting with, or being affected by AI technologies. In stigmatized healthcare settings such as sexual health and HIV prevention, comfort may be especially relevant because individuals must disclose highly personal information, navigate privacy and judgment concerns, and engage with technologies in emotionally sensitive contexts.

Despite growing interest in AI-enabled HIV prevention, evidence describing how women perceive these tools is limited. Evidence from adjacent healthcare domains suggests that concerns about privacy, bias, transparency, misdiagnosis, data security, and loss of human connection may reduce trust, comfort, and adoption.^21^ Conversely, technology familiarity, prior digital health use, and perceived usefulness may increase openness to AI-enabled care.^22^ To our knowledge, no validated measure has been developed specifically to assess comfort with AI-enabled tools in HIV prevention or sexual healthcare. Moreover, attitudes toward AI are unlikely to be homogeneous across populations. Identifying distinct AI knowledge and comfort profiles among women may inform implementation strategies, communication approaches, and patient-centered design of future HIV prevention technologies.

In this study, we aimed to 1) characterize latent subgroups of women based on self-reported AI knowledge and comfort; 2) describe how these subgroups differed across sociodemographic and behavioral characteristics and healthcare technology utilization; and 3) examine whether and the degree to which subgroup membership was associated with PrEP awareness.

## Methodology

### Study Design and Setting

We conducted a cross-sectional survey of English and Spanish-speaking adults aged 18 and older residing in the greater New York City (NYC) area between August 2023 and August 2024. Participants were recruited online through social media sites (Facebook, Instagram, Amazon Turk), and Columbia University’s recruitment website (https://recruit.cumc.columbia.edu/), and in-person through posted flyers and community engagement in the catchment area of NewYork-Presbyterian/Columbia University Irving Medical Center (NYP/CUIMC), a large quaternary academic medical system. Recruitment used a nonprobability sampling design.

All participants provided informed consent and completed a one-time, self-administered electronic survey hosted on REDCap, a secure web platform.^23^ Each respondent received a pre-loaded debit card worth $50 as a survey incentive. The analytic sample comprised all eligible respondents accrued during the study period. The questionnaire assessed individual and household-level social determinants of health (SDoH), medical history, sexual and drug use risk and preventative behaviors, and health technology utilization. An additional module on artificial intelligence adapted from a national survey developed by Khullar and colleagues was administered to participants completing the questionnaire after April 25, 2024, following human subjects approval.^21^ Full item wording, response options, and coding for all measures and outcomes used in the analysis are provided as supplementary materials (Table S1). This study was approved by the CUIMC Institutional Review Board (Protocol: AAAT6079). Reporting adheres to the Strengthening the Reporting of Observational Studies in Epidemiology (STROBE) statement for cross-sectional studies.^24^

### Participants

For this study, the analytic sample comprised all eligible respondents accrued during the study period who self-identified as women, were aged 18 years or older, resided in the greater New York City area, completed all AI-related questions in survey version 2, and reported never having received a formal diagnosis of HIV (and thus might benefit from PrEP).

### Conceptual Model

We developed a conceptual model grounded in theories of trust, comfort, and technology acceptance that conceptualizes AI comfort as a multidimensional construct reflecting both willingness to engage with AI under uncertainty and affective ease with AI-enabled interactions (**Figure 1**).^25–28^ AI comfort included six sub-domains and 11 indicators: self-reported level of AI knowledge (one item), comfort contributing anonymous personal data for AI training (one item), comfort sharing private information with an AI chatbot (one item), comfort in sharing private information with healthcare providers (one item), comfort with AI-supported decisions about HIV or STI testing (two items), and comfort with transparency and validity of AI-supported diagnosis or treatment decisions at varying levels of transparency and validity (five items).

**Figure 1:**
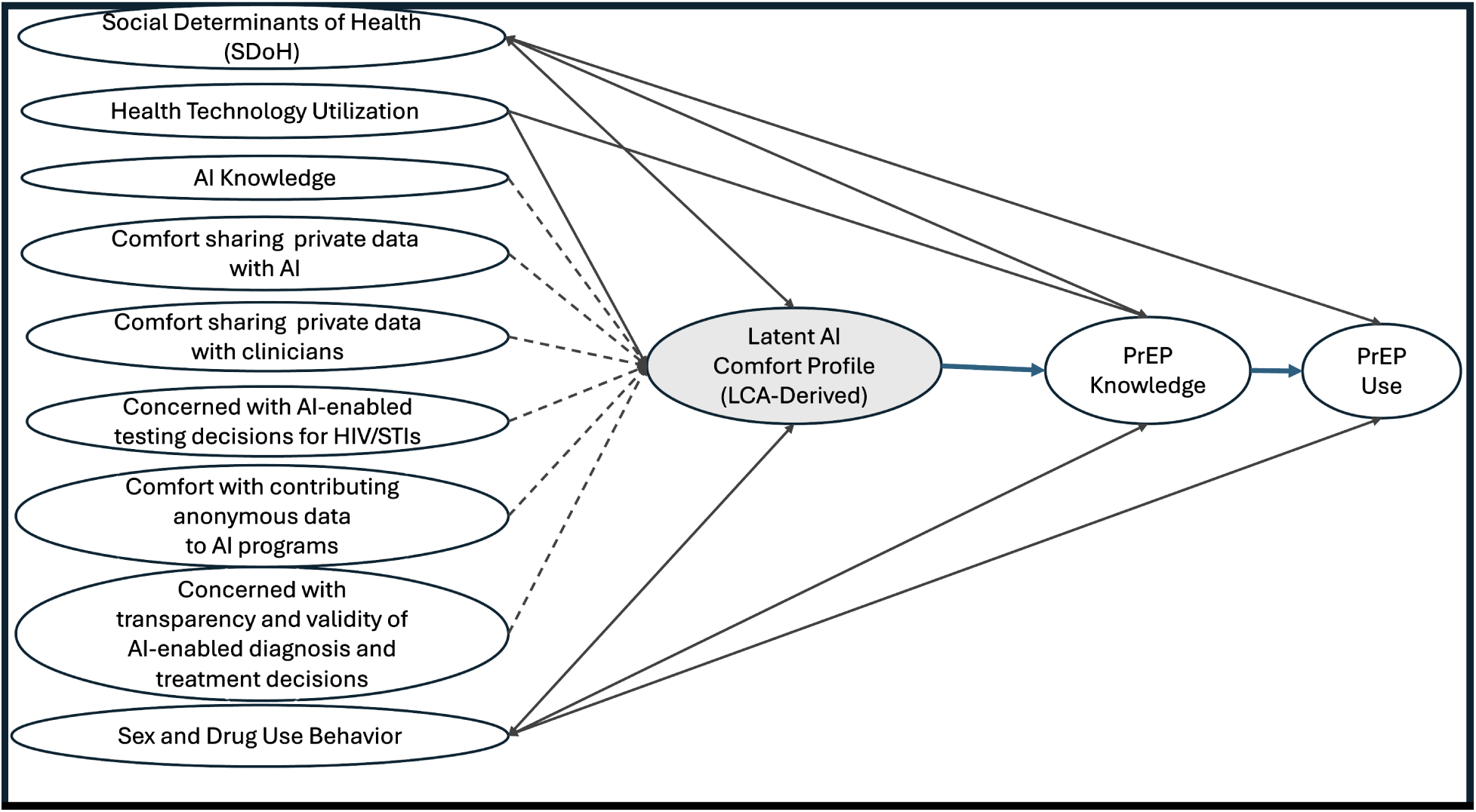
Conceptual model of Latent AI comfort Profiles and PrEP Knowledge and Use. Latent AI comfort is conceptualized as a multidimensional construct and operationalized by six constructs (dashed lines); self-assessed level of AI knowledge, comfort sharing private information with an AI-enabled tool like a chatbot, comfort sharing private information with clinicians, comfort with AI-supported HIV/STI testing decisions, comfort with sharing anonymous data to improve AI programs, and concerns with transparency and validity of AI-enabled diagnosis and treatment decisions. Healthcare technology utilization, social determinants of health (SDoH), and HIV-associated behaviors are hypothesized to confound or modify the relationship between latent profiles of comfort with AI-enabled tools and PrEP awareness and use. Pathways represent hypothesized conceptual relationships rather than causal effects.

In healthcare contexts, willingness may be shaped both by perceived technical performance and how AI systems are implemented within patient-clinician relationships.^25–28^ Consistent with implementation-oriented frameworks, the model further posits that health technology utilization, which may reflect familiarity and engagement with digital tools, and SDoH and sexual and drug use behavior, which may influence healthcare access, risk perception, and exposure to HIV prevention information, may also affect AI comfort and PrEP-related outcomes through individual, system-level, and contextual pathways.^22^ Given the cross-sectional nature of the study, the pathways shown in **Figure 1** represent hypothesized conceptual relationships rather than causal relationships.

### Measures and Outcomes

The primary outcome was self-reported HIV PrEP awareness, dichotomized responses as “Yes” versus “No /Not sure” based on the question, “Have you heard of pre-exposure prophylaxis (PrEP), a medication (pill or injections) to prevent getting HIV?” The secondary outcome, PrEP use, was classified with response options “Yes” or “No” based on the question, “Do you currently take PrEP to prevent getting HIV?”

### Primary Exposure

The primary exposure, AI comfort, was operationalized as latent class membership derived from eleven indicators representing six sub-domains as described above in the conceptual model. Full item wording, response options, and coding are provided in **Table S1**. Indicators were recoded as four-point ordinal responses, with lower values indicating lower concern or greater comfort and higher values indicating greater concern, lower comfort, or uncertainty.

### Other Measures

#### Health Technology Utilization

The composite healthcare technology utilization construct was comprised of eight indicators described in detail in **Table S1**. Each indicator was re-coded as binary: “yes=1” and “no/ I don’t know=0” and summed for a total composite score ranging from 0-8. In brief, the composite indicator of health technology utilization was comprised of the following variables: ownership or use of tablet or smartphone, mobile banking, medical needs, use of tablets or smartphones to make decision, and the use of health portals to communicate with a provider in the last 12 months.

#### SDoH and HIV-Associated Behaviors

Sociodemographic covariates included age, race and ethnicity, language, educational attainment, employment status, and borough or residential location. Sexual and drug use behavior variables included perceived HIV and STI risk, condom use, number of partners in the last 90 days, and prior drug use.

### Latent Class Analysis

We fit the latent mixture model using the six recoded AI indicators. We compared latent class analysis (LCA) models between 2 and 7 classes after 5000 bootstrap resampling using the following model fit statistics: log likelihood, Akaike information criterion (AIC), Bayesian information criterion (BIC), entropy, and Chi-squared values. The final class solution was selected based on overall model fit, parsimony, and interpretability in relation to the conceptual framework. Participants were assigned to the class for which they had the highest posterior probability. Classes were characterized by indicators with the highest posterior probabilities representing AI knowledge and comfort.

### Statistical Analysis

For multi-item domains/sub-domains, we computed Cronbach’s alpha, with *α≥*0.70 for internal consistency and as the threshold for composite score construction. Participant characteristics were summarized overall and by PrEP awareness status, PrEP use, and latent classes using frequencies and percentages for categorical values and means (*±* SD) or medians (IQR) for continuous variables, as appropriate. We assessed bivariate comparisons using Chi-squared or Fisher’s exact tests for categorical variables and Student’s t-tests or Wilcoxon rank-sum tests for continuous variables, depending on variable distribution and expected cell counts. We estimated associations between latent profiles and PrEP awareness using logistic regression to generate unadjusted and adjusted odds ratios with 95% confidence intervals. We adjusted for potential confounding variables based on the conceptual framework and statistical associations with both latent class and PrEP awareness. Statistical significance was assessed using two-sided tests with *α*=0.05. All analyses were conducted using R (Version 4.5.3; R Core Team, 2026).

### Sensitivity Analyses

All variables had complete responses, reducing bias due to missingness. The primary outcome (PrEP knowledge), “No” and “Don’t know” was combined, so we explored misclassification by examining the separated responses. We also fit alternative regression models that adjusted for variables associated with PrEP awareness but was not associated with latent profiles. Sensitivity analyses were conducted for other explanatory variables that were re-categorized due to small cell frequencies.

## Results

### Participant Characteristics

Among 2,745 survey respondents, 306 cisgender women met inclusion criteria for the analytic sample. The median age was 36 years (IQR, 27–46), and most participants were aged 25–49 years (64%). Participants were mostly Hispanic (60%), followed by non-Hispanic Black (18%), non-Hispanic White (12%), and other/multi-race (10%); English-speaking or bilingual in English and Spanish (81%), had not completed college (53%), and were employed (58%). Participants resided in the Bronx (32%), followed by Manhattan (18%), Brooklyn (17%), Queens (16%), and other NYC metropolitan areas (17%) (**Table 1; Table S2**).

**Table 1:**
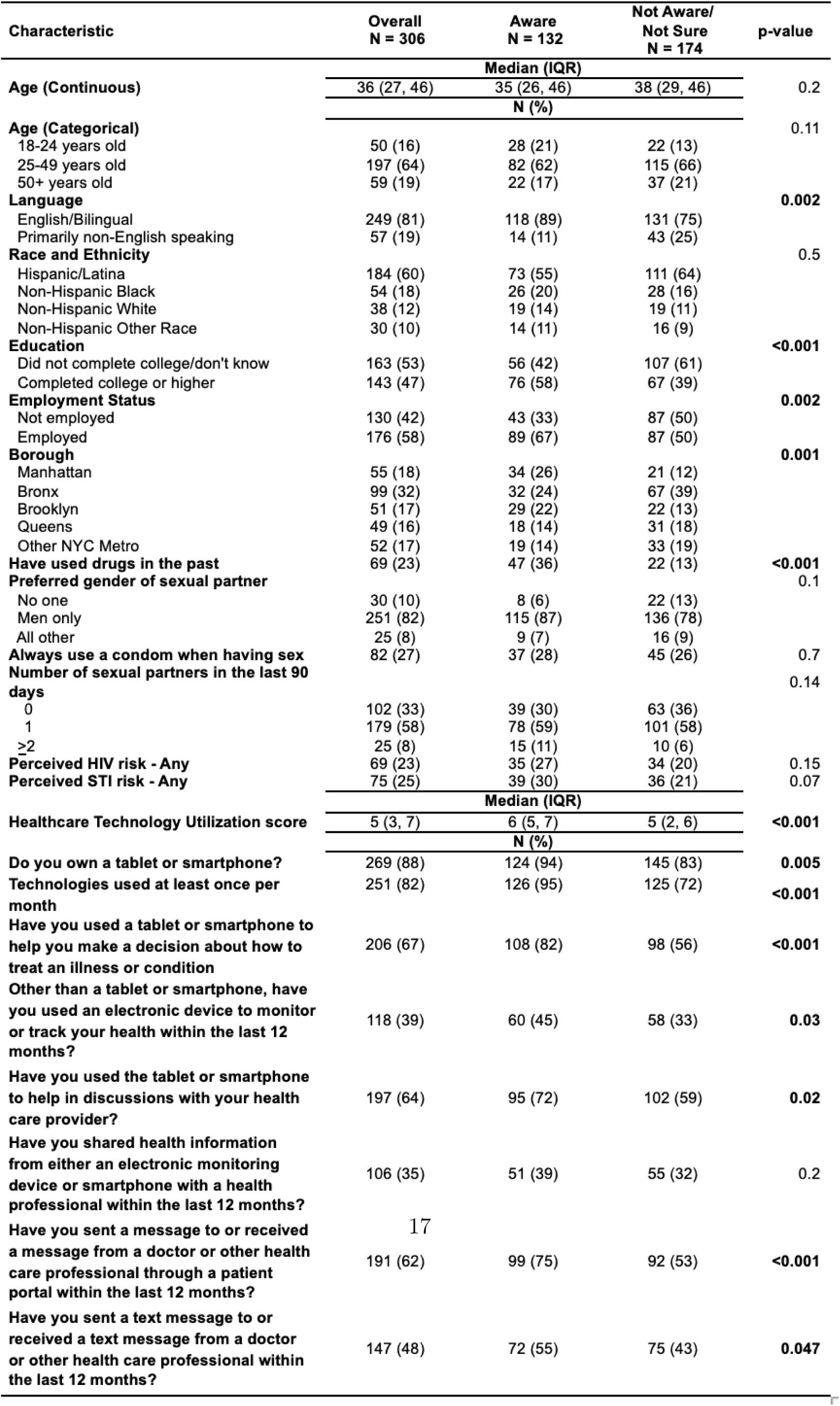
Sociodemographic Characteristics, Sexual and Drug Use Behaviors Overall and by PrEP Awareness Status Among Women in the New York City Metropolitan Area.

Overall, 132 participants (43%) reported being aware of PrEP and 12 (4%) reported current PrEP use. Due to small frequencies, we only report summary statistics regarding PrEP use (**Table S3**). **Table 1** shows the distribution overall and by PrEP awareness. PrEP awareness was more prevalent among English-speaking or bilingual than Spanish-speaking only (89% vs 75%; p=0.002), those who completed college or higher (58%) versus less than college (39%; p<0.001), and employed versus unemployed women (67% vs 50%; p=0.002). PrEP awareness also differed by borough of residence (p=0.001), with a higher proportion of PrEP-aware participants residing in Manhattan (62%) or Brooklyn (57%) compared to Queens (37%), other metro areas (37%), or the Bronx (32%). Prior drug use was more common among PrEP-aware participants (36% vs 13%; p<0.001). Condom use, number of sexual partners in the past 90 days, perceived HIV or STI risk did not differ by PrEP awareness. We observed the same differences when PrEP awareness was disaggregated “no” and “don’t know” (**Table S2**). Additionally, differences were observed for gender of sexual partners, and perceived HIV and STI risk perception.

### AI Knowledge and Comfort

Two-thirds of participants reported little or no knowledge of AI. Concerns about AI-supported healthcare were also common. Most participants (66%) were somewhat or very concerned that AI could make an incorrect diagnosis, and nearly half were somewhat or very uncomfortable with an AI algorithm being used to determine whether they needed HIV screening. At the same time, responses suggested conditional openness to AI-enabled tools when privacy protections were explicit: 57% reported being somewhat or very comfortable sharing information with an AI chatbot if they believed the information would remain confidential, and 39% believed AI would make healthcare somewhat or much better in the next five years. Among multi-item constructs, healthcare technology utilization showed good internal consistency (Cronbach’s *α*=0.80), as did AI-supported test decision-making (*α*=0.89) and AI-supported diagnosis or treatment (*α*=0.74). These constructs were therefore summarized using composite scores and included as indicators in the latent class analysis.

### Healthcare Technology Utilization

Healthcare technology utilization was common: 88% owned a tablet or smartphone, 82% used technologies at least monthly, 64% used a tablet or smartphone to communicate with a healthcare provider, and 62% used patient portal messaging in the prior 12 months (**Table 1**). The median healthcare technology utilization score was 5 (IQR: 3–7). PrEP-aware participants had a higher healthcare technology utilization score than unaware or unsure participants (median 6 [IQR, 5–7] vs 5 [IQR, 2–6]; p<0.001).

### Latent AI Knowledge and Comfort Profiles

Latent class model fit statistics supported a four-class solution as the optimal balance of model fit, class separation, and parsimony based on log likelihood, AIC, BIC, entropy, and substantive interpretability (**Table 2**). Model fit improved as the number of classes increased, as reflected by decreasing AIC values. The four-class model demonstrated high entropy (0.90), indicating good classification accuracy, and provided the best balance of model fit, parsimony, class size, and interpretability. Although the five-class model had similar entropy, it had a higher BIC and was less parsimonious. The four-class model was therefore selected as the final solution.

**Table 2:**
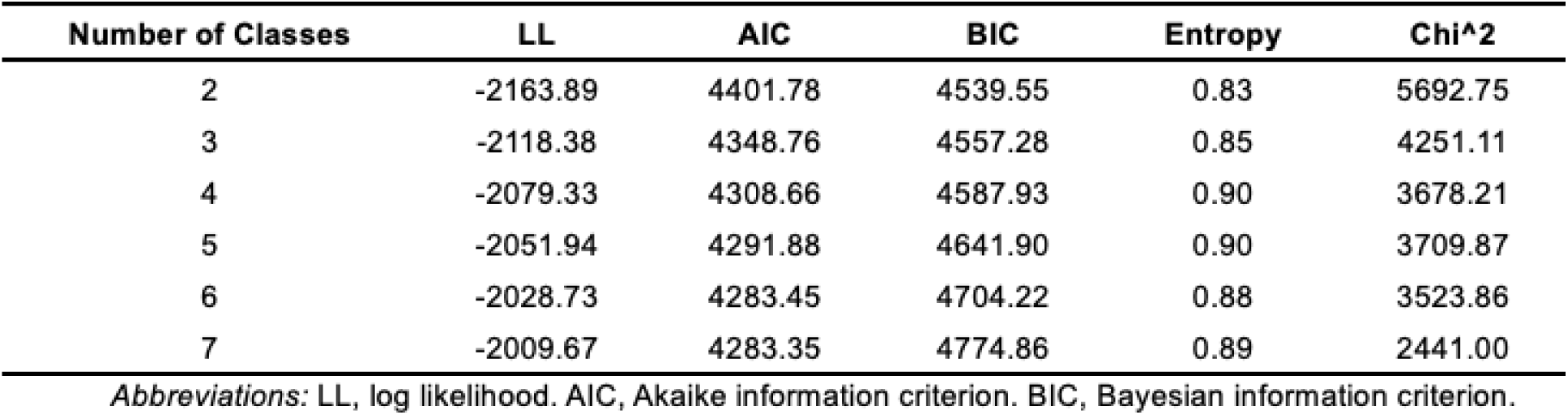
Fit Statistics for Latent Class Analysis (2-7 classes).

The four-class solution identified distinct AI knowledge and comfort profiles (**Table 3**; **Figure 2**). Class 1 (uninformed and cautiously comfortable) included 47 participants (15%) and was characterized by low or no AI knowledge, uncertainty about selected aspects of AI use, but relative comfort with privacy protections when sharing information with either AI chatbots or human clinicians. Class 2 (uninformed and selectively uncomfortable) was the largest group, composed of 154 participants (50%), and was characterized by low AI knowledge and high uncertainty across multiple aspects of AI use, including uncertainty about both AI-mediated and human-provider interactions. Class 3 (uninformed and broadly uncomfortable) included 29 participants (9%) and was characterized by varied AI knowledge but broad discomfort with AI, including discomfort with chatbot privacy, while retaining comfort with human clinicians maintaining privacy. Class 4 (somewhat informed and selectively comfortable) included 76 participants (25%) and was characterized by moderate AI knowledge and selective comfort with AI, including comfort with chatbot privacy and similar comfort with human clinicians maintaining privacy.

**Figure 2:**
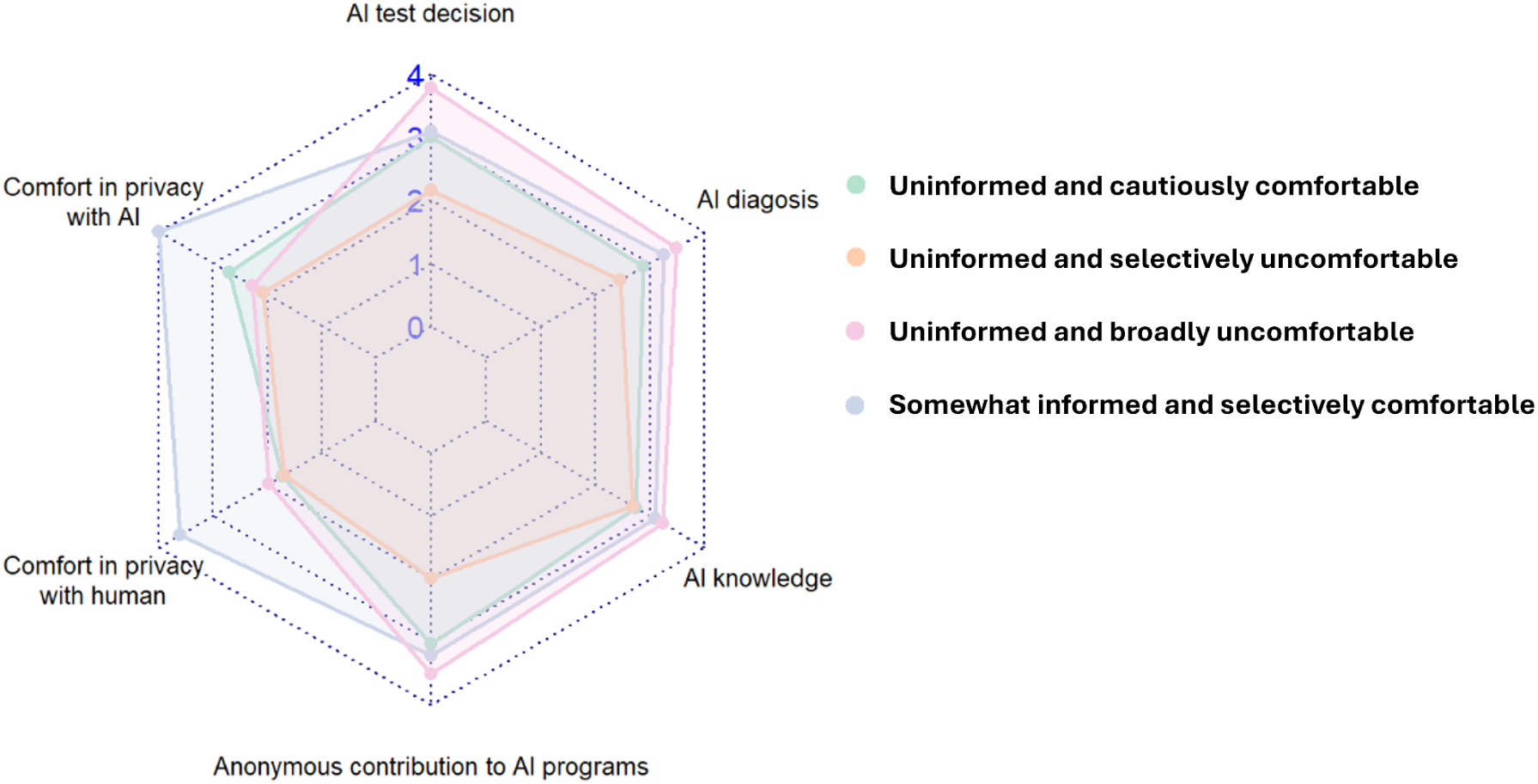
Radar Plot of Latent AI Knowledge and Comfort Profiles Among Women in the New York City Metropolitan Area (n=306).

**Table 3:**
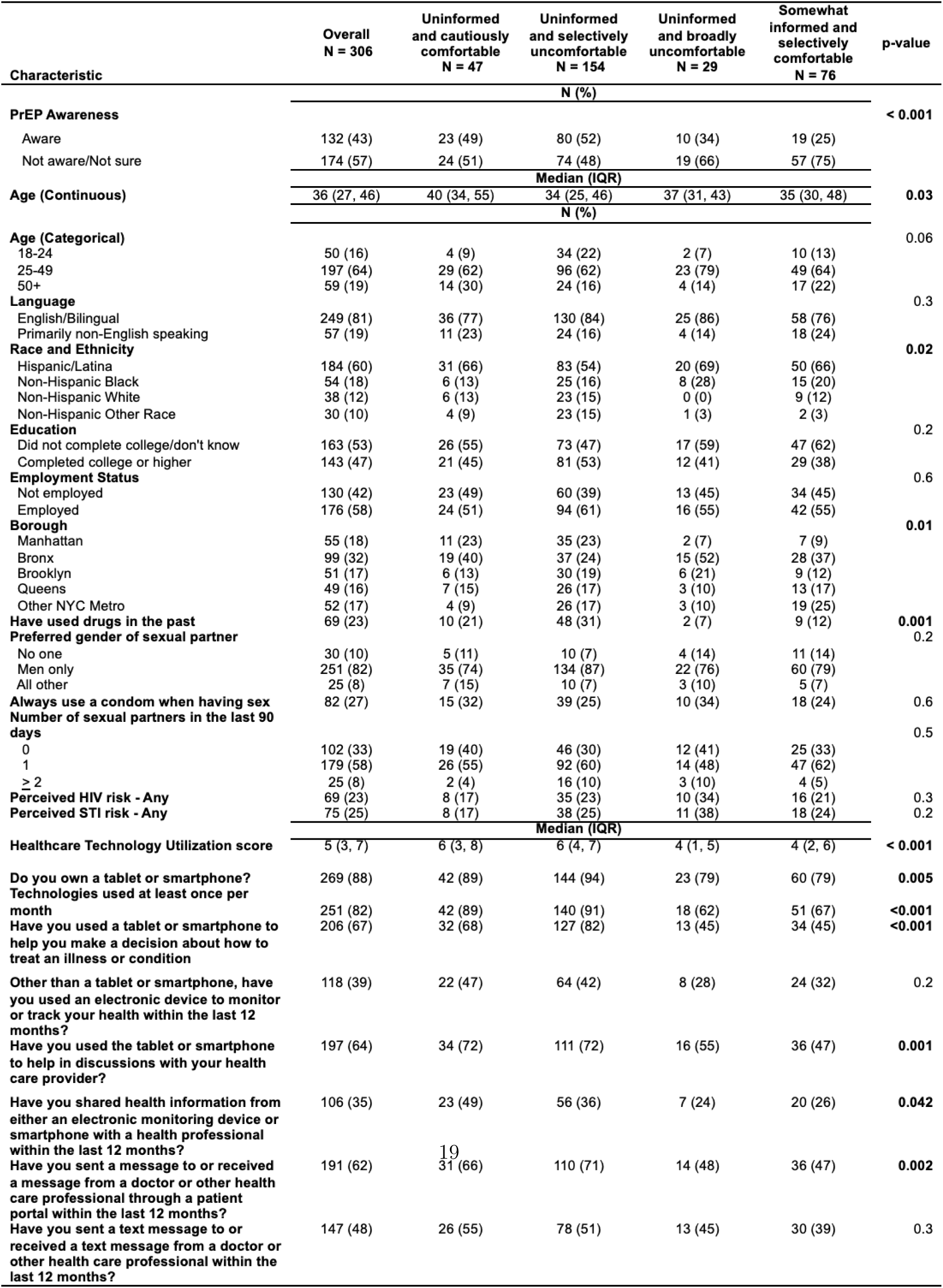
Sociodemographic Characteristics, Sexual and Drug Use Behaviors Overall and by Latent Classes Among Women in the New York City Metropolitan Area.

Latent class membership differed significantly by several participant characteristics. Race and ethnicity varied across classes (p=0.02), as did borough of residence (p=0.01). Participants who rated themselves as uninformed and broadly uncomfortable with AI were disproportionately Hispanic or Black and more likely to reside in the Bronx, while Manhattan residents were more concentrated among respondents who were uninformed and either cautiously or selectively comfortable with AI. Latent class membership also differed by prior drug use (p=0.001) and healthcare technology utilization (p<0.001). Participants in latent classes comprising uninformed individuals who were either cautiously or selectively comfortable with AI had higher median healthcare technology utilization scores and higher levels of monthly technology use, smartphone or tablet ownership, use of devices in healthcare decision-making, provider communication, and patient portal messaging than others.

### Association Between AI Knowledge/Comfort Profiles and PrEP Awareness

In unadjusted analyses, PrEP awareness differed by latent AI knowledge/comfort profile (**Table 4**). Compared with women who rated themselves as uninformed and cautiously comfortable with AI, participants who were somewhat informed selectively uncomfortable with AI had significantly lower odds of being aware of PrEP (OR, 0.35; 95% CI, 0.16–0.75; p=0.007) while uninformed participants selectively comfortable with AI had similar odds of PrEP awareness (OR, 1.13; 95% CI, 0.59–2.18; p=0.7) as did participants who were uninformed and broadly comfortable had lower odds (OR, 0.55; 95% CI, 0.21–1.41; p=0.2). After adjusting for healthcare technology utilization, and borough of residence, there was no statistical association between latent AI knowledge/comfort profile and PrEP awareness.

**Table 4:** Association of Latent AI Knowledge and Comfort Profiles and PrEP Awareness Among Women in the New York City Metropolitan Area.

| Characteristic | Overall<br>N = 306 | Unaware/<br>Not Sure<br>N = 174 | Aware<br>N = 132 |  | Unadjusted<br>Odds Ratio |  | Adjusted<br>Odds Ratio<sup>a</sup> |  |
| --- | --- | --- | --- | --- | --- | --- | --- | --- |
|  |  | N (%) |  | p-value | OR (95% CI) | p-value | aOR (95% CI) | p-value |
| **Latent AI Knowledge/Comfort Profile** |  |  |  | **<0.001** |  |  |  |  |
| Uninformed and cautiously comfortable | 47 (15) | 24 (51) | 23 (49) |  | Reference |  | Reference |  |
| Uninformed and selectively uncomfortable | 154 (50) | 74 (48) | 80 (52) |  | 1.13 (0.59-2.18) | 0.7 | 0.96 (0.47-1.93) | 0.9 |
| Uninformed and broadly uncomfortable | 29 (9) | 19 (66) | 10 (34) |  | 0.55 (0.21-1.41) | 0.2 | 0.49 (0.21-1.12) | 0.09 |
| Somewhat informed and selectively comfortable | 76 (25) | 57 (75) | 19 (25) |  | 0.35 (0.16-0.75) | 0.007 | 0.85 (0.30-2.35) | 0.8 |
| **Borough** |  |  |  | **0.001** |  |  |  |  |
| Manhattan | 55 (18) | 21 (12) | 34 (26) |  | Reference |  | Reference |  |
| Bronx | 99 (32) | 67 (39) | 32 (24) |  | 0.29 (0.15-0.58) | **<0.001** | 0.39 (0.18-0.81) | **0.01** |
| Brooklyn | 51 (17) | 22 (13) | 29 (22) |  | 0.81 (0.37-1.77) | 0.6 | 0.92 (0.40-2.10) | 0.8 |
| Queens | 49 (16) | 31 (18) | 18 (14) |  | 0.36 (0.16-0.79) | **0.01** | 0.38 (0.16-0.87) | **0.03** |
| Other NYC Metro | 52 (17) | 33 (19) | 19 (14) |  | 0.36 (0.16-0.77) | **0.01** | 0.53 (0.22-1.21) | 0.1 |
| **Have used drugs in the past** | 69 (23) | 22 (13) | 47 (36) | **<0.001** | 3.82 (2.18-6.87) | **<0.001** | 2.80 (1.53-5.23) | **<0.001** |
|  |  | Median (IQR) |  |  | OR (95% CI) | p-value | aOR (95% CI) | p-value |
| **Healthcare technology utilization score** | 5 (3, 7) | 5 (2, 6) | 6 (5, 7) | **<0.001** | 1.28 (1.15-1.43) | **<0.001** | 1.2 (1.06-1.35) | **0.003** |

<sup>a</sup>Fully adjusted model accounts for confounding due to differences in healthcare technology utilization and borough of residence, covariates identified from the conceptual framework that were statistically significant in bivariate comparisons.

Several covariates remained associated with PrEP awareness after statistical adjustment (**Table S4**). Compared with participants residing in Manhattan, participants residing in the Bronx had lower adjusted odds of PrEP awareness (aOR, 0.46; 95% CI, 0.21–0.97; p=0.04), as did participants residing in Queens (aOR, 0.37; 95% CI, 0.16–0.86; p=0.02). Prior drug use was associated with higher adjusted odds of PrEP awareness (aOR, 2.39; 95% CI, 1.29–4.51; p=0.006). Greater healthcare technology utilization was also associated with higher adjusted odds of PrEP awareness (aOR per one-point increase, 1.19; 95% CI, 1.05–1.34; p=0.006). After adjustment, statistical associations with PrEP awareness did not persist for language, education, or employment status, or Brooklyn or other NYC metropolitan area residence.

## Discussion

In this cross-sectional study of cisgender women in New York City who were potentially eligible for PrEP, we found limited self-reported AI knowledge, and substantial heterogeneity in comfort with AI-enabled healthcare applications. Latent class analysis identified four distinct AI knowledge/comfort profiles that differed by PrEP awareness before statistical adjustment. The association between AI profiles and PrEP awareness did not persist after adjustment, suggesting that social, geographic, behavioral, and technology-related factors explained observed differences. AI comfort profiles also differed by race/ethnicity, borough of residence, prior drug use, and healthcare technology utilization in PrEP awareness across AI comfort profiles. These findings also illustrate the importance of distinguishing AI familiarity from AI comfort.^29,30^ AI comfort appeared task-specific: participants expressed greater concern about AI-supported diagnosis and testing decisions than about privacy-protected information sharing. This distinction is particularly relevant for HIV prevention, where stigma, confidentiality, trust, and patient-clinician communication influence engagement with care.

PrEP awareness was also low and varied by participants’ AI knowledge/comfort profiles suggesting that AI readiness and HIV prevention awareness may intersect in socially patterned ways. Participants with profiles characterized by greater AI discomfort or selective comfort had lower PrEP awareness and lower healthcare technology utilization.^31^ These findings suggest that comfort with AI may represent broader differences (e.g., digital inclusion, healthcare access, prior healthcare experiences, and exposure to HIV prevention information).^32^ AI-enabled HIV prevention tools should account for the contexts in which women encounter both HIV prevention services and digital health technologies.

These findings also have design and implementation implications for AI-enabled PrEP education and HIV prevention tools.^33^ Many participants were comfortable sharing information with an AI chatbot when confidentiality was assured, suggesting that transparent, privacy-preserving applications may be acceptable to some women. However, discomfort with AI-supported diagnosis and testing decisions suggests that early applications in sexual health may be better positioned as supportive, educational, or navigational tools rather than autonomous clinical decision-makers.

### Limitations

This study has several limitations in study design, measurement, and inference. The cross-sectional design precludes conclusions about temporal ordering or causality. All measures were self-reported and may be subject to recall bias, social desirability bias, or differing interpretation. Latent class measured comfort, but did not measure trust, which is closely related but distinct. Some latent class frequencies were relatively small, limiting statistical power. Participants were recruited using a nonprobability sampling approach from an urban area, which also may limit generalizability. Finally, we measured PrEP awareness rather than knowledge that PrEP is available for women, which may provide greater insights for implementation. The prevalence of current PrEP use was too low to conduct inferential analyses. Future studies should examine how AI comfort relates to more detailed outcomes across the PrEP continuum. Despite these limitations, the study provides early evidence on how women perceive AI-enabled tools in the context of HIV prevention. By identifying distinct AI knowledge and comfort profiles and examining their relationship to PrEP awareness, this analysis contributes to a growing evidence base on patient-centered, equity-oriented implementation of AI in sexual health.

## Conclusions

Among cisgender women in New York City, limited PrEP awareness and limited AI knowledge were common, but many participants expressed conditional openness to AI-enabled tools when confidentiality was assured. AI knowledge/comfort profiles differed by PrEP awareness and by social, geographic, behavioral, and technology-related factors, although adjusted analyses suggested that these relationships may be partly explained by broader contextual factors, particularly borough of residence. These findings suggest AI-enabled HIV prevention tools should be designed for technical accuracy as well as trust, transparency, confidentiality, and the lived contexts of the women they intend to serve.

## Data Availability

All data produced in the present study are available upon reasonable request to the authors.

## Acknowledgements

We wish to thank study participants for their time and the members of our Community Advisory Board for their invaluable input throughout the course of the study.

## Data availability statement

The data that support the findings of this study are available from the corresponding author (DC) upon request.

## Institutional Review Board Statement

The study was conducted in accordance with the Declaration of Helsinki and approved by the Institutional Review Board of Columbia University Irving Medical Center (AAAT6079).

## Informed Consent Statement

Informed consent was obtained from all subjects involved in the study.

## Author Contributions

Dr. Castor had full access to the data in the study and takes responsibility for the integrity of the data and accuracy of the analysis. All authors have approved the final manuscript.

Conceptualization: AFN, DC, HRN, MES, MF, DT

Funding: DC, MES

Investigation: All authors Methodology: DC, HRN

Statistical Analysis: DC, HRN, MF, SH

Supervision: DC, MES

Visualization: DC, HRN, MF, SH

Writing – Original Draft: DC, HRN, MF

Writing – Review & Editing: All authors

---

**Figure S1:**
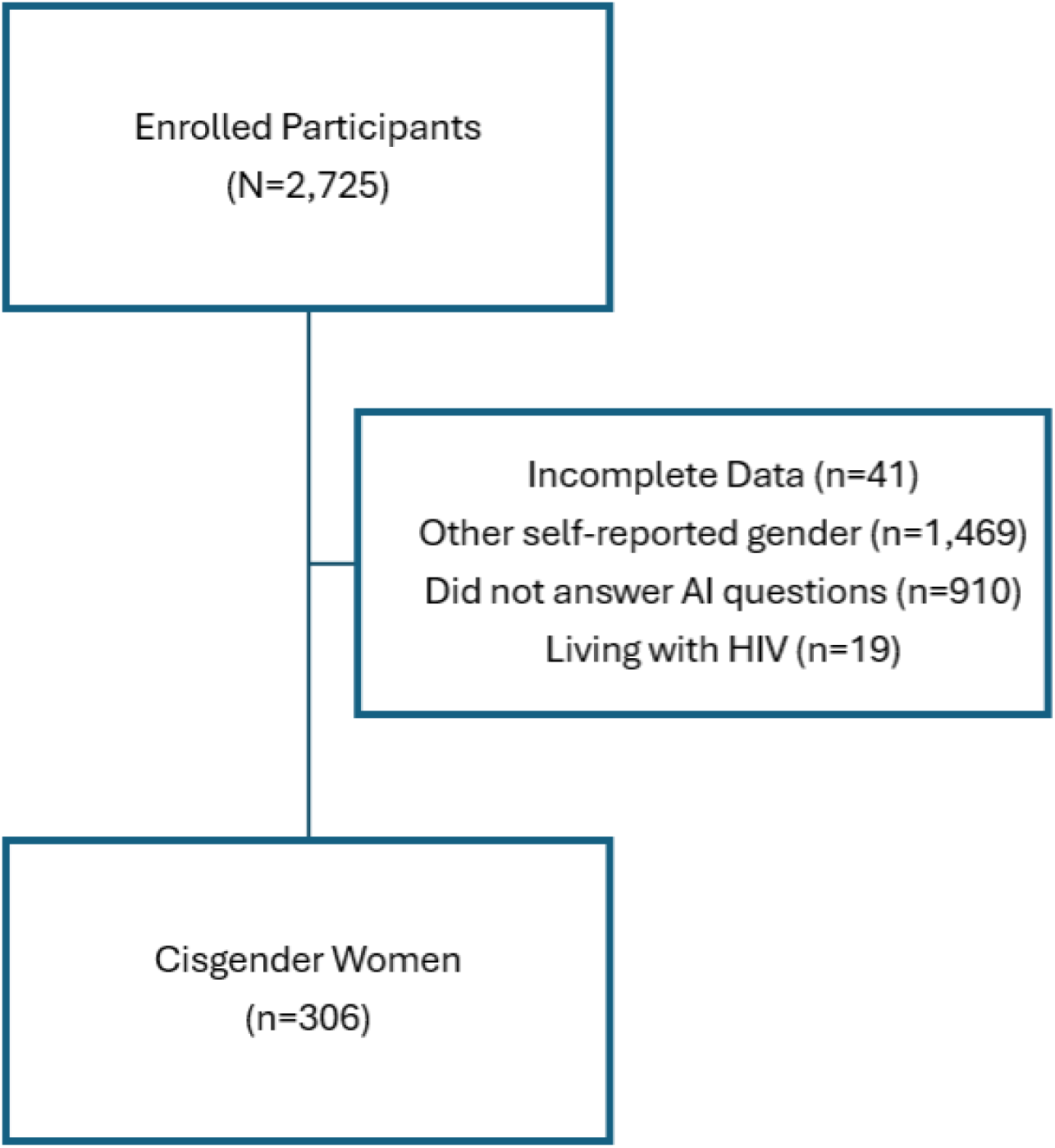
Participant Flow Diagram.

**Table S1:**
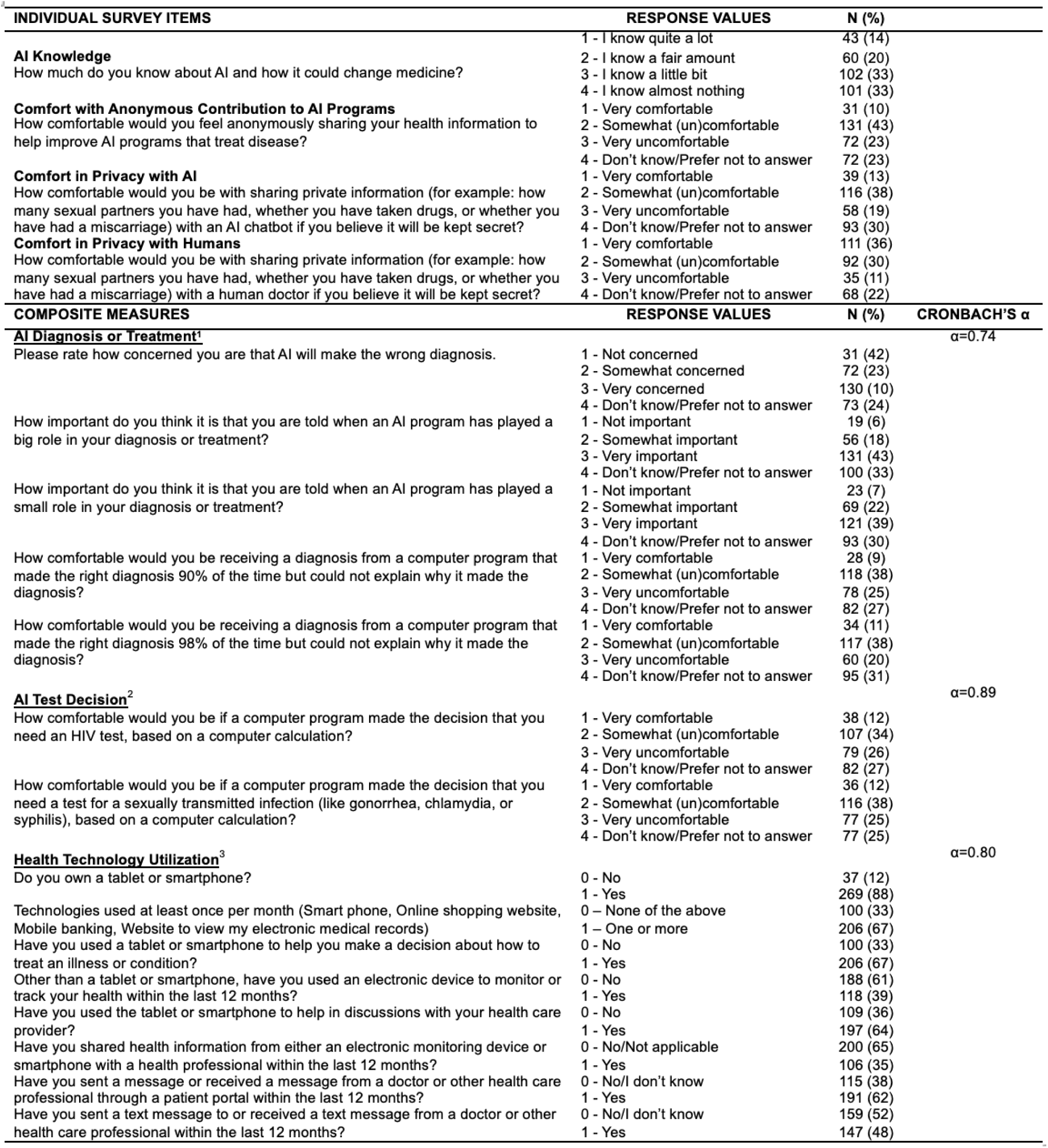
Item wording, response options, and coding for study measures and outcomes.

**Table S2:** Sociodemographic Characteristics, Sexual and Drug Use Behaviors Overall and by PrEP Awareness Status (Aware, Not Aware, Not Sure) Among Women in the New York City Metropolitan Area.

| Characteristic | Overall<br>N = 306 | Aware<br>N = 132 | Not Aware<br>N = 174 | Not Sure<br>N = 46 | p-value |
| --- | --- | --- | --- | --- | --- |
| **Median (IQR)** |  |  |  |  |  |
| **Age (Continuous)** | 36 (27, 46) | 35 (26, 46) | 38 (29, 48) | 37 (27, 45) | 0.3 |
| **N (%)** |  |  |  |  |  |
| **Age (Categorical)** |  |  |  |  | 0.2 |
| 18-24 years old | 50 (16) | 28 (21) | 14 (11) | 8 (17) |  |
| 25-49 years old | 197 (64) | 82 (62) | 86 (67) | 29 (63) |  |
| 50+ years old | 59 (19) | 22 (17) | 28 (22) | 9 (20) |  |
| **Language** |  |  |  |  | 0.007 |
| English/Bilingual | 249 (81) | 118 (89) | 97 (76) | 34 (74) |  |
| Primarily non-English speaking | 57 (19) | 14 (11) | 31 (24) | 12 (26) |  |
| **Race and Ethnicity** |  |  |  |  | 0.5 |
| Hispanic/Latina | 184 (60) | 73 (55) | 78 (61) | 33 (72) |  |
| Non-Hispanic Black | 54 (18) | 26 (20) | 20 (16) | 8 (17) |  |
| Non-Hispanic White | 38 (12) | 19 (14) | 17 (13) | 2 (4) |  |
| Non-Hispanic Other Race | 30 (10) | 14 (11) | 13 (10) | 3 (7) |  |
| **Education** |  |  |  |  | <0.001 |
| Did not complete college/don't know | 163 (53) | 56 (42) | 70 (55) | 37 (80) |  |
| Completed college or higher | 143 (47) | 76 (58) | 58 (45) | 9 (20) |  |
| **Employment Status** |  |  |  |  | 0.001 |
| Not employed | 130 (42) | 43 (33) | 58 (45) | 29 (63) |  |
| Employed | 176 (58) | 89 (67) | 70 (55) | 17 (37) |  |
| **Borough** |  |  |  |  | <0.001 |
| Manhattan | 55 (18) | 34 (26) | 17 (13) | 4 (9) |  |
| Bronx | 99 (32) | 32 (24) | 38 (30) | 29 (63) |  |
| Brooklyn | 51 (17) | 29 (22) | 16 (13) | 6 (13) |  |
| Queens | 49 (16) | 18 (14) | 30 (23) | 1 (2) |  |
| Other NYC Metro | 52 (17) | 19 (14) | 27 (21) | 6 (13) |  |
| **Have used drugs in the past** | 69 (23) | 47 (36) | 19 (15) | 3 (7) | <0.001 |
| **Preferred gender of sexual partner** |  |  |  |  | 0.048 |
| No one | 30 (10) | 8 (6) | 12 (9) | 10 (22) |  |
| Men only | 251 (82) | 115 (87) | 104 (81) | 32 (70) |  |
| All other | 25 (8) | 9 (7) | 12 (9) | 4 (9) |  |
| **Always use a condom when having sex** | 82 (27) | 37 (28) | 31 (24) | 14 (30) | 0.7 |
| **Number of sexual partners in the last 90 days** |  |  |  |  | 0.2 |
| 0 | 102 (33) | 39 (30) | 43 (34) | 20 (43) |  |
| 1 | 179 (58) | 78 (59) | 78 (61) | 23 (50) |  |
| ≥2 | 25 (8) | 15 (11) | 7 (6) | 3 (7) |  |
| **Perceived HIV risk - Any** | 69 (23) | 35 (27) | 16 (13) | 18 (39) | <0.001 |
| **Perceived STI risk - Any** | 75 (25) | 39 (30) | 15 (12) | 21 (46) | <0.001 |
| **Median (IQR)** |  |  |  |  |  |
| **Healthcare Technology Utilization score** | 5 (3, 7) | 6 (5, 7) | 5 (3, 7) | 4 (1, 5) | < 0.001 |
| **N (%)** |  |  |  |  |  |
| **Do you own a tablet or smartphone?** | 269 (88) | 124 (94) | 112 (88) | 33 (72) | <0.001 |
| **Technologies used at least once per month** | 251 (82) | 126 (95) | 100 (78) | 25 (54) | <0.001 |
| **Have you used a tablet or smartphone to help you make a decision about how to treat an illness or condition** | 206 (67) | 108 (82) | 81 (63) | 17 (37) | <0.001 |
| **Other than a tablet or smartphone, have you used an electronic device to monitor or track your health within the last 12 months?** | 118 (39) | 60 (45) | 44 (34) | 14 (30) | 0.09 |
| **Have you used the tablet or smartphone to help in discussions with your health care provider?** | 197 (64) | 95 (72) | 79 (62) | 23 (50) | 0.02 |
| **Have you shared health information from either an electronic monitoring device or smartphone with a health professional within the last 12 months?** | 106 (35) | 51 (39) | 44 (34) | 11 (24) | 0.2 |
| **Have you sent a message to or received a message from a doctor or other health care professional through a patient portal within the last 12 months?** | 191 (62) | 99 (75) | 76 (59) | 16 (35) | <0.001 |
| **Have you sent a text message to or received a text message from a doctor or other health care professional within the last 12 months?** | 147 (48) | 72 (55) | 63 (49) | 12 (26) | 0.004 |

**Table S3:**
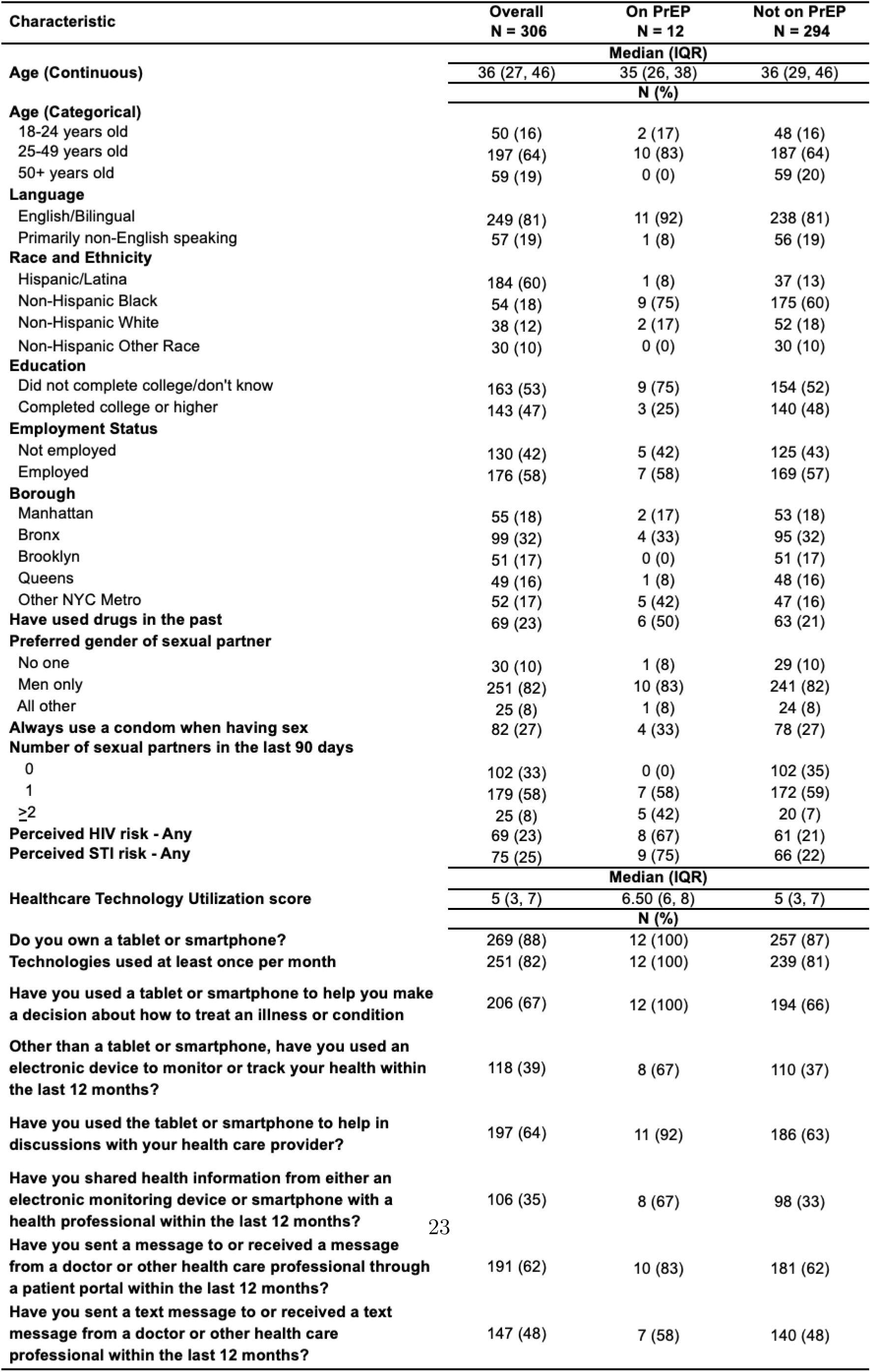
Sociodemographic Characteristics, Sexual and Drug Use Behaviors Overall and by PrEP Use Status Among Women in the New York City Metropolitan Area.

| Characteristic | Overall<br>N = 306 | On PrEP<br>N = 12 | Not on PrEP<br>N = 294 |
| --- | --- | --- | --- |
|  | **Median (IQR)** |  |  |
| **Age (Continuous)** | 36 (27, 46) | 35 (26, 38) | 36 (29, 46) |
|  | **N (%)** |  |  |
| **Age (Categorical)** |  |  |  |
| 18-24 years old | 50 (16) | 2 (17) | 48 (16) |
| 25-49 years old | 197 (64) | 10 (83) | 187 (64) |
| 50+ years old | 59 (19) | 0 (0) | 59 (20) |
| **Language** |  |  |  |
| English/Bilingual | 249 (81) | 11 (92) | 238 (81) |
| Primarily non-English speaking | 57 (19) | 1 (8) | 56 (19) |
| **Race and Ethnicity** |  |  |  |
| Hispanic/Latina | 184 (60) | 1 (8) | 37 (13) |
| Non-Hispanic Black | 54 (18) | 9 (75) | 175 (60) |
| Non-Hispanic White | 38 (12) | 2 (17) | 52 (18) |
| Non-Hispanic Other Race | 30 (10) | 0 (0) | 30 (10) |
| **Education** |  |  |  |
| Did not complete college/don't know | 163 (53) | 9 (75) | 154 (52) |
| Completed college or higher | 143 (47) | 3 (25) | 140 (48) |
| **Employment Status** |  |  |  |
| Not employed | 130 (42) | 5 (42) | 125 (43) |
| Employed | 176 (58) | 7 (58) | 169 (57) |
| **Borough** |  |  |  |
| Manhattan | 55 (18) | 2 (17) | 53 (18) |
| Bronx | 99 (32) | 4 (33) | 95 (32) |
| Brooklyn | 51 (17) | 0 (0) | 51 (17) |
| Queens | 49 (16) | 1 (8) | 48 (16) |
| Other NYC Metro | 52 (17) | 5 (42) | 47 (16) |
| **Have used drugs in the past** | 69 (23) | 6 (50) | 63 (21) |
| **Preferred gender of sexual partner** |  |  |  |
| No one | 30 (10) | 1 (8) | 29 (10) |
| Men only | 251 (82) | 10 (83) | 241 (82) |
| All other | 25 (8) | 1 (8) | 24 (8) |
| **Always use a condom when having sex** | 82 (27) | 4 (33) | 78 (27) |
| **Number of sexual partners in the last 90 days** |  |  |  |
| 0 | 102 (33) | 0 (0) | 102 (35) |
| 1 | 179 (58) | 7 (58) | 172 (59) |
| ≥2 | 25 (8) | 5 (42) | 20 (7) |
| **Perceived HIV risk - Any** | 69 (23) | 8 (67) | 61 (21) |
| **Perceived STI risk - Any** | 75 (25) | 9 (75) | 66 (22) |
|  | **Median (IQR)** |  |  |
| **Healthcare Technology Utilization score** | 5 (3, 7) | 6.50 (6, 8) | 5 (3, 7) |
|  | **N (%)** |  |  |
| **Do you own a tablet or smartphone?** | 269 (88) | 12 (100) | 257 (87) |
| **Technologies used at least once per month** | 251 (82) | 12 (100) | 239 (81) |
| **Have you used a tablet or smartphone to help you make a decision about how to treat an illness or condition** | 206 (67) | 12 (100) | 194 (66) |
| **Other than a tablet or smartphone, have you used an electronic device to monitor or track your health within the last 12 months?** | 118 (39) | 8 (67) | 110 (37) |
| **Have you used the tablet or smartphone to help in discussions with your health care provider?** | 197 (64) | 11 (92) | 186 (63) |
| **Have you shared health information from either an electronic monitoring device or smartphone with a health professional within the last 12 months?** | 106 (35) | 8 (67) | 98 (33) |
| **Have you sent a message to or received a message from a doctor or other health care professional through a patient portal within the last 12 months?** | 191 (62) | 10 (83) | 181 (62) |
| **Have you sent a text message to or received a text message from a doctor or other health care professional within the last 12 months?** | 147 (48) | 7 (58) | 140 (48) |

**Table S4:** Association of Latent AI Knowledge and Comfort Profiles and PrEP Awareness Among Women in the New York City Metropolitan Area.

| Characteristic | Overall<br>N = 306 | Unaware/<br>Not Sure<br>N = 174 | Aware<br>N = 132 |  | Unadjusted<br>Odds Ratio | Adjusted Odds Ratio<sup>a</sup> |  |  |
| --- | --- | --- | --- | --- | --- | --- | --- | --- |
|  |  | N (%) |  | p-value | OR (95% CI) | p-value | aOR (95% CI) | p-value |
| **Latent AI Knowledge/Comfort Profile** |  |  |  | **< 0.001** |  |  |  |  |
| Uninformed and cautiously comfortable | 47 (15) | 24 (51) | 23 (49) |  | Reference |  | Reference |  |
| Uninformed and selectively uncomfortable | 154 (50) | 74 (48) | 80 (52) |  | 1.13 (0.59, 2.18) | 0.7 | 0.90 (0.44, 1.84) | 0.8 |
| Uninformed and selectively uncomfortable | 29 (9) | 19 (66) | 10 (34) |  | 0.55 (0.21, 1.41) | 0.2 | 0.74 (0.26, 2.07) | 0.6 |
| Somewhat informed and selectively comfortable | 76 (25) | 57 (75) | 19 (25) |  | 0.35 (0.16, 0.75) | **0.007** | 0.46 (0.19, 1.06) | 0.07 |
| **Language** |  |  |  | **0.002** |  |  |  |  |
| English/Bilingual | 249 (81) | 131 (75) | 118 (89) |  | Reference |  | Reference |  |
| Primarily non-English speaking | 57 (19) | 43 (25) | 14 (11) |  | 0.36 (0.18, 0.68) | **0.002** | 0.54 (0.25, 1.11) | 0.1 |
| **Education** |  |  |  | **< 0.001** |  |  |  |  |
| Did not complete college/don't know | 163 (53) | 107 (61) | 56 (42) |  | Reference |  | Reference |  |
| Completed college or higher | 143 (47) | 67 (39) | 76 (58) |  | 2.17 (1.37, 3.45) | **0.001** | 1.37 (0.80, 2.36) | 0.3 |
| **Employment Status** |  |  |  | **0.002** |  |  |  |  |
| Not employed | 130 (42) | 87 (50) | 43 (33) |  | Reference |  | Reference |  |
| Employed | 176 (58) | 87 (50) | 89 (67) |  | 2.07 (1.30, 3.33) | **0.002** | 1.34 (0.75, 2.38) | 0.3 |
| **Borough** |  |  |  | **0.001** |  |  |  |  |
| Manhattan | 55 (18) | 21 (12) | 34 (26) |  | Reference |  | Reference |  |
| Bronx | 99 (32) | 67 (39) | 32 (24) |  | 0.29 (0.15, 0.58) | **<0.001** | 0.46 (0.21, 0.97) | **0.04** |
| Brooklyn | 51 (17) | 22 (13) | 29 (22) |  | 0.81 (0.37, 1.77) | 0.6 | 0.82 (0.35, 1.93) | 0.7 |
| Queens | 49 (16) | 31 (18) | 18 (14) |  | 0.36 (0.16, 0.79) | **0.01** | 0.37 (0.16, 0.86) | **0.02** |
| Other NYC Metro | 52 (17) | 33 (19) | 19 (14) |  | 0.36 (0.16, 0.77) | **0.01** | 0.52 (0.21, 1.23) | 0.14 |
| **Have used drugs in the past** | 69 (23) | 22 (13) | 47 (36) | **<0.001** | 3.82 (2.18, 6.87) | **<0.001** | 2.39 (1.29, 4.51) | **0.006** |
|  |  | Median (IQR) |  |  | OR (95% CI) | p-value | aOR (95% CI) | p-value |
| **Healthcare technology utilization score** | 5 (3, 7) | 5 (2, 6) | 6 (5, 7) | **<0.001** | 1.28 (1.15, 1.43) | **<0.001** | 1.19 (1.05, 1.34) | **0.006** |

<sup>a</sup>Fully adjusted model accounts for variables associated with PrEP awareness (i.e., language, education, employment status, borough of residence, prior drug use, and healthcare technology utilization score).

